# Clinical and demographic factors modify the association between plasma phosphorylated tau-181 and cognition

**DOI:** 10.1101/2023.11.03.23298051

**Authors:** Corey J. Bolton, Marilyn Steinbach, Omair A. Khan, Dandan Liu, Julia O’Malley, Logan Dumitrescu, Amalia Peterson, Angela L. Jefferson, Timothy J. Hohman, Henrik Zetterberg, Katherine A. Gifford, Alzheimer’s Disease Neuroimaging Initiative

## Abstract

**INTRODUCTION:** Plasma phosphorylated tau181 (p-tau181) associations with global cognition and memory are clear, but the link between p-tau181 with other cognitive domains and subjective cognitive decline (SCD) across the clinical spectrum of Alzheimer’s disease (AD) and how this association changes based on genetic and demographic factors is poorly understood.

**METHODS:** Participants were drawn from the Alzheimer’s Disease Neuroimaging Initiative and included 1185 adults aged >55 years with plasma p-tau181 and neuropsychological test data. Linear regression models related plasma p-tau181 to neuropsychological composite and SCD scores with follow-up models examining plasma p-tau181 interactions with cognitive diagnosis, *APOE* ε4 carrier status, age, and sex on cognitive outcomes.

**RESULTS:** Higher plasma p-tau181 was associated with worse memory, executive functioning, and language abilities, and greater informant-reported SCD. Visuospatial abilities and self-report SCD were not associated with plasma p-tau181. Associations were generally stronger in MCI or dementia, *APOE* ε4 carriers, women, and younger participants.

**DISCUSSION:** Higher levels of plasma p-tau181 are associated with worse neuropsychological test performance across multiple cognitive domains; however, these associations vary based on disease stage, genetic risk status, age, and sex.

## Introduction

Advancements in Alzheimer’s disease (AD) therapies have necessitated early and accurate detection of AD pathology to initiate treatment prior to widespread neuronal loss. Blood-based biomarkers for phosphorylated tau (p-tau) have emerged as accessible and specific biomarkers for AD pathology. Plasma levels of tau phosphorylated at threonine 181 (plasma p-tau181) are highly correlated with cerebrospinal fluid levels of p-tau181,^1^ associated with amyloid and tau deposition on positron emission tomography imaging,^2,3^ relate to neurodegeneration in AD-specific brain regions on volumetric imaging,^4^ and accurately differentiate AD from other neurodegenerative conditions.^1,3^ Due to its promise in identifying AD pathological changes, this biomarker has potential for integration into clinical settings for the screening and diagnosis of AD.^5^

While plasma p-tau181 appears to reflect the underlying pathology of AD, its associations with clinical outcomes are less understood. Past work has identified associations between plasma p-tau181 on cognitive screening measures or isolated measures of memory functioning.^3,4^ However, there has been little work examining more subtle changes in cognition or patterns of cognitive decline across neuropsychological domains. Identifying domains of cognition (e.g., episodic memory, executive functioning) that are affected can provide valuable information related to localization of pathological changes, thereby further elucidating the clinicopathological correlates of this novel biomarker.^6^ Further, it is not yet known how associations between plasma p-tau181 and cognition may be modified by demographic data (e.g., age, sex), disease state (mild cognitive impairment (MCI), dementia) or genetic factors (e.g., *apolipoprotein E (APOE)-*ε4 carrier status) which are known to be associated with cognition in aging.^7–9^

This study investigates associations between plasma p-tau181 and comprehensive assessment of objective and subjective cognition. Results will elucidate how this novel biomarker relates to specific clinical changes across various disease states and demographic groups. We hypothesize that plasma p-tau181 will be most strongly associated with memory and executive functions, consistent with a typical AD clinical syndrome,^10^ with stronger associations in individuals at increased risk of clinical AD (e.g., *APOE* ε4 carriers, individuals with MCI).

## Methods

### Participants

Data used in the preparation of this article were obtained from the Alzheimer’s Disease Neuroimaging Initiative (ADNI) database (adni.loni.usc.edu). The ADNI was launched in 2003 as a public-private partnership, led by Principal Investigator Michael W. Weiner, MD. The primary goal of ADNI has been to test whether serial magnetic resonance imaging (MRI), positron emission tomography (PET), other biological markers, and clinical and neuropsychological assessment can be combined to measure the progression of MCI and AD. Data were obtained from the ADNI database (https://www.loni.ucla.edu/ADNI/Data/ADCS_Download.jsp) in November 2021 and included participants from cohorts ranging from ADNI 1 to ADNI 3. Participants aged 55-90 were recruited from across the United States and Canada and were excluded if they had a Hachinski ischemic score^11^ >4, Geriatric Depression Scale (GDS)^12^ score ≥6, no study partner, contraindications for MRI, did not speak English or Spanish fluently, were not in good physical health, or used drugs with anticholinergic or opioid properties. Cognitively unimpaired (CU) participants had no memory complaints, normal memory function (assessed using delayed paragraph recall of the Wechsler Memory Scale–Revised; WMS-R)^13^ and Mini Mental State Examination (MMSE)^14^ ≥24, Clinical Dementia Rating (CDR)^15^ global score of 0, and absence of significant impairment in cognitive and functional performance. Participants with mild cognitive impairment (MCI) had memory complaint (by self-report or from study partner), abnormal memory functioning (>1 SD below the normative sample on WMS-R) and MMSE ≥24, CDR global score of 0.5, and sufficient cognitive and functional performance so as to prevent diagnosis of dementia. Participants with dementia had memory complaint, abnormal memory function and MMSE between 20 and 26, CDR global score of 0.5-1.0, and NINCDS/ADRDA criteria for probable AD.^16^ For more information, see www.adni-info.org. Participants without usable plasma p-tau181 or covariate data were excluded.

### Neuropsychological Measures

Participants completed comprehensive neuropsychological assessment of memory, executive functioning, language, and visuospatial abilities using the ADNI neuropsychological protocol. Domain-based composite scores were utilized. These composite scores were previously developed using item response theory and latent variable modeling to minimize the possibility of multiple comparisons from similar assessments. The memory composite^17^ included the Rey Auditory Verbal Learning Test (RAVLT),^18^ ADAS-Cog^19^ word list and word recognition, WMS-R Logical Memory,^13^ and MMSE^14^ word recall. The executive functioning composite^20^ included Category Fluency,^21^ Trails A and B,^22^ Digit Span Backwards,^23^ Digit Symbol Substitution,^23^ and Clock Drawing.^24^ The visuospatial composite^25^ included Clock Drawing, the interlocking pentagons of the MMSE,^14^ and the constructional praxis item on ADAS-Cog. The language composite included the Boston Naming Test;^26^ Category Fluency; the MMSE’s object naming, sentence repetition, reading, and writing, and following a three step command; ADAS-Cog commands, object naming, and ideational praxis; and the Montreal Cognitive Assessment’s phonemic fluency and sentence repetition.^27^ Single-factor models constructed using Mplus or R were used to create each composite; models were fit to the data, which was assessed using confirmatory fit index, Tucker-Lewis index, and root mean squared error of approximation. These models were completed by ADNI and resulting composite scores were downloaded. Higher composite scores indicate better test performance.

### Subjective Cognitive Decline

Participants and their loved ones completed the Everyday Cognition (ECog) questionnaire as a measure of self- and informant-reported subjective cognitive decline (SCD). The 39-item ECog questionnaire assesses functional abilities linked to different cognitive domains (memory, language, and executive functioning) compared to 10 years prior using a 5-point scale with higher scores indicating greater SCD.^28^

### Fluid Collection and Biochemical Analyses

Participants completed a fasting venous blood draw. Plasma p-tau181 concentration was measured using Single molecule array (Simoa) technology, using an in-house assay developed by the Clinical Neurochemistry Laboratory, University of Gothenburg, Sweden, described fully elsewhere.^3^

### Statistical Analyses

Linear regression models with ordinary least square estimates related plasma p-tau181 to objective and subjective cognition outcomes. Covariates were selected *a priori* based on their potential to confound analytical models due to their known associations with objective and subjective cognition, including age,^29^ sex,^30^ education^31^, cognitive diagnosis,^32^ and *APOE* ε4 carrier status (defined as having at least one *APOE* ε4 allele).^33^ To account for changes in p-tau181 levels in different clinical and demographic groups, subsequent models evaluated *plasma p-tau181 x cognitive diagnosis, p-tau181 x APOE ε4 carrier status, p-tau181 x age,* and *p-tau181 x sex* interactions on objective and subjective cognition. Significance was set *a priori* at *p* < 0.05. Sensitivity analyses were performed by removing outliers >4 standard deviations from mean values to assess whether outliers accounted for results. Additional sensitivity analyses were conducted for ECog outcomes using a mean score rather than a sum of scores to determine if score quantification accounted for results. Multiple comparison correction was performed for each analytical set using a false discovery rate based on the Benjamini-Hochberg procedure.^34^ Analyses were conducted using R 3.4.2 (www.r-project.org).

## Results

Participants included 1185 adults ages 55 to 94 (46% female, 44% *APOE* ε4 carriers). See **Table 1** for participant characteristics for the entire sample and stratified by cognitive diagnosis. Participants were also divided by *APOE* ε4 carrier status, age (using a median split), and sex for stratified analyses. *APOE* ε4 carriers, compared to noncarriers, were younger, more likely to have MCI or dementia, performed worse on memory, executive functions, and language composite measures, had higher p-tau181 levels, and had greater self- and informant-reported SCD. Compared to the younger age group, the older group were more likely to be male, less likely to be *APOE* ε4 carriers, more likely to have dementia over MCI, performed worse on memory, executive functions, and language composite measures, had more p-tau181, and had greater informant-reported SCD. Women, compared to men, were more likely to be CU, had less education, were younger, performed better on memory composite, had lower p-tau181, and had lower self- and informant-reported SCD.

**Table 1.**
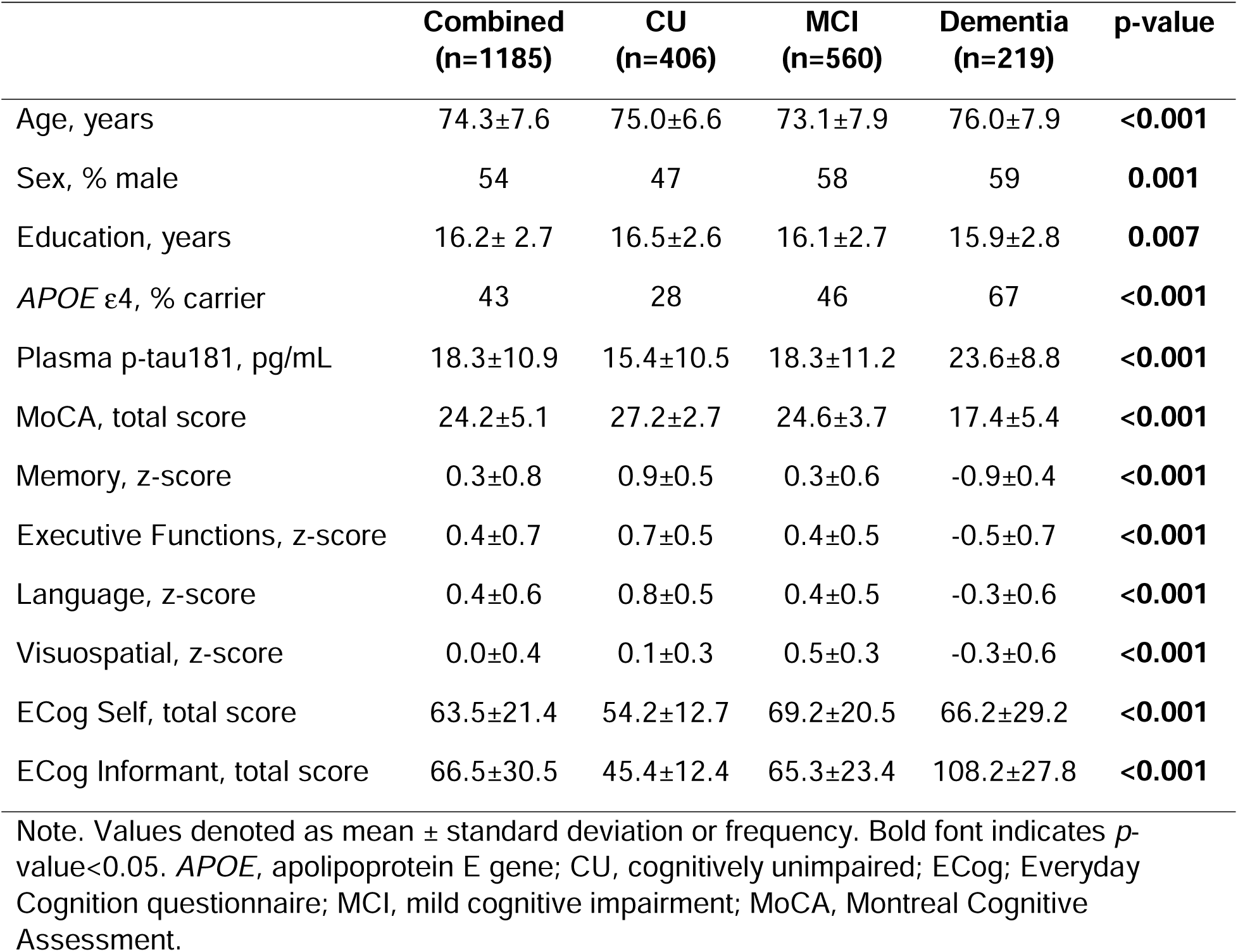
Participant Characteristics

### Plasma P-tau181 and Cognition

In the entire sample, higher p-tau181 levels were associated with lower scores on memory (β=-0.008, p-value<0.001), executive functioning (β=-0.005, p-value=0.002), and language (β=-0.004, p-value=0.006) composite scores, but not on visuospatial functioning (β=-0.001, p-value=0.47). Additionally, higher p-tau181 levels were associated with higher informant-reported SCD (β=0.17, p-value=0.007) but not with self-reported SCD (β=0.07, p-value=0.59). Results were comparable after FDR correction. See **Table 2** for results of main effects analyses.

**Table 2.** Plasma P-Tau181 Associations with Cognitive and SCD Outcomes

| | $\beta$ | 95% Confidence Intervals | *p*-value |
| --- | --- | --- | --- |
| Memory | -0.008 | -0.01, -0.005 | **<0.001\*** |
| Executive Functions | -0.005 | -0.008, -0.002 | **0.002\*** |
| Language | -0.004 | -0.007, -0.001 | **0.008\*** |
| Visuospatial | -0.001 | -0.004, 0.002 | 0.47 |
| ECog Self | 0.07 | -0.18, 0.32 | 0.59 |
| ECog Informant | 0.17 | 0.05, 0.29 | **0.007\*** |

Note: Models were adjusted for age, sex, education, *APOE* ε4 status, and cognitive diagnosis. Bold font indicates *p*-value<0.05. \*FDR-corrected *p*-value<0.05. *APOE*, apolipoprotein E gene; CU, cognitively unimpaired; ECog, Everyday Cognition questionnaire; MCI, mild cognitive impairment.

### Plasma P-tau181 x Cognitive Diagnosis and Cognition

P-tau181 interacted with diagnosis on memory (β=-0.001, p-value<0.001), executive functioning (β=-0.01, p-value=0.01), language (β=-0.009, p-value=0.05), and informant-report SCD (β=-0.51, p-value=0.03), but not on visuospatial composite score or self-reported SCD (*p*-values>0.12). Models stratified by diagnosis revealed these associations were driven by MCI and dementia participants. Higher p-tau181 was associated with lower performance on memory (β=-0.01, p-value<0.001), executive functioning (β=-0.005, p-value=0.01), and language (β=-0.008, p-value=0.02) composites in participants with MCI as well as lower executive functioning (β=-0.03, p-value=0.008) and language (β=-0.02, p-value=0.03) composite scores and higher informant-reported SCD (β=0.52, p-value=0.02) scores in participants with dementia. Results were comparable after FDR correction. See **Table 3** for results stratified by diagnosis and **Figure 1** for illustrations of stratified analyses.

**Table 3.** Plasma P-Tau181 x Diagnosis Interactions on Cognitive and SCD Outcomes

|  | *p-tau x diagnosis* |  | CU (n=406) |  | MCI (n=560) |  | Dementia (n=219) |  |
| --- | --- | --- | --- | --- | --- | --- | --- | --- |
| | $\beta$ | *p*-value | $\beta$ | *p*-value | $\beta$ | *p*-value | $\beta$ | *p*-value |
| Memory | 0.001 | **<0.001\*** | -0.002 | 0.36 | -0.01 | **<0.001\*** | -0.002 | 0.47 |
| Executive Functions | -0.01 | **0.01\*** | 0.001 | 0.55 | -0.005 | **0.01\*** | -0.02 | **0.008\*** |
| Language | -0.009 | **0.05** | 0.0002 | 0.94 | -0.004 | **0.02\*** | -0.01 | **0.03** |
| Visuospatial | -0.003 | 0.75 | 0.001 | 0.54 | -0.001 | 0.41 | -0.002 | 0.68 |
| ECog Self | -0.53 | 0.13 | 0.10 | 0.37 | 0.14 | 0.43 | -0.23 | 0.61 |
| ECog Informant | 0.51 | **0.03** | 0.009 | 0.90 | 0.18 | 0.07 | 0.52 | **0.02\*** |

Note: Models were adjusted for age, sex, education, and *APOE* ε4 status. Bold font indicates *p*-value<0.05. \*FDR-corrected *p*-value<0.05. *APOE*, apolipoprotein E gene; CU, cognitively unimpaired; ECog, Everyday Cognition questionnaire; MCI, mild cognitive impairment.

**Figure 1.**
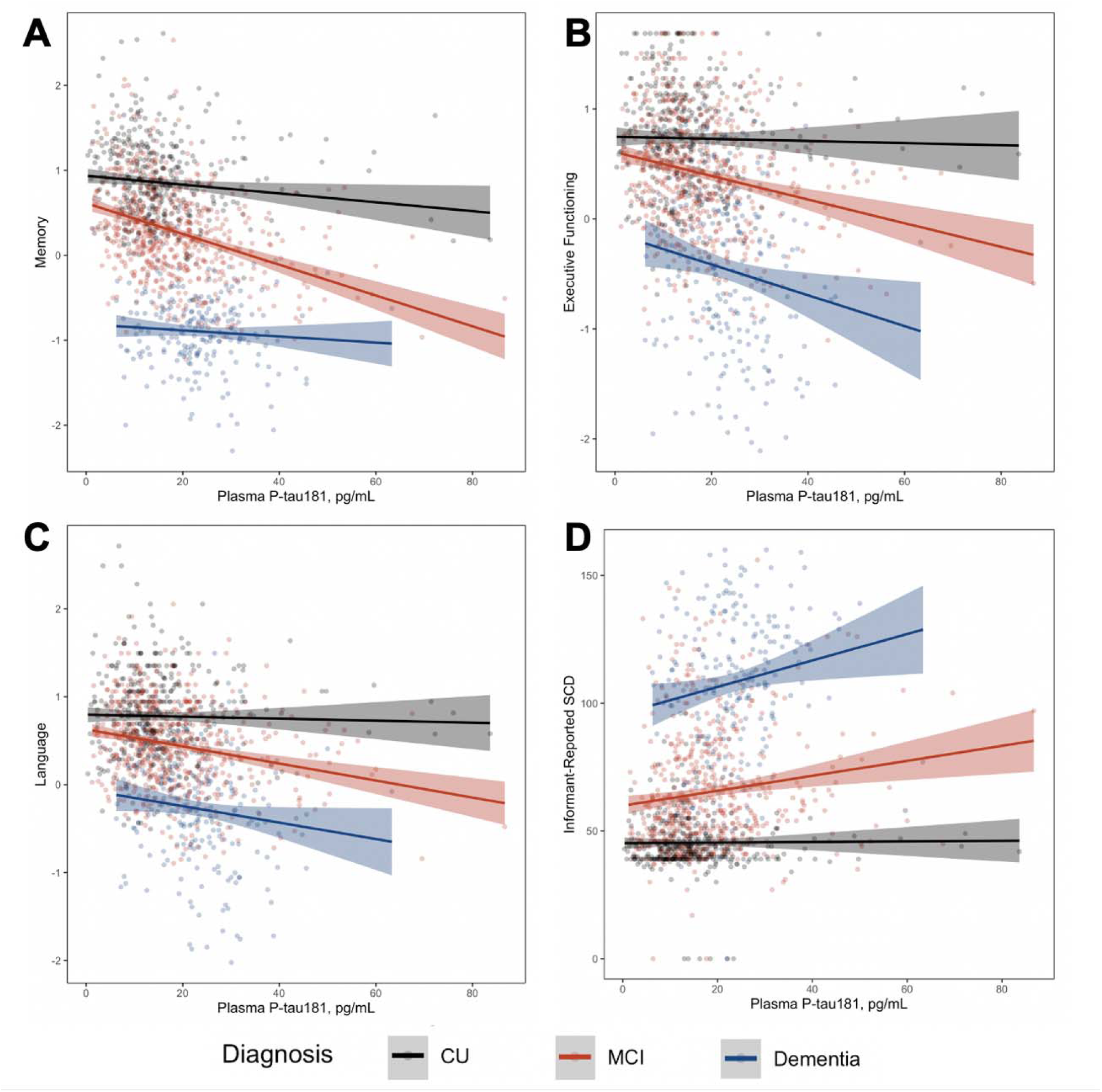
Plasma p-tau181 x Cognitive Diagnosis Interactions on Cognitive Outcomes. Lines reflect cognitive domain or informant-reported SCD scores corresponding to plasma p-tau181 levels. Shading reflects 95% confidence interval. (A) Associations between plasma p-tau181 levels and memory composite score, stratified by cognitive diagnosis; CU β=-0.002, p=0.36; MCI β=-0.01, p<0.001; Dementia β=-0.002, p=0.47. (B) Associations between plasma p-tau181 levels and executive functioning composite score, stratified by cognitive diagnosis; CU β=0.001, p=0.55; MCI β=-0.005, p=0.01; Dementia β=-0.02, p=0.008. (C) Associations between plasma p-tau181 levels and language composite score, stratified by cognitive diagnosis; CU β=0.0002, p=0.94; MCI β=-0.004, p=0.02; Dementia β=-0.01, p=0.03. (D) Associations between plasma p-tau181 levels and informant-reported SCD score, stratified by cognitive diagnosis; CU β=0.0009, p=0.90; MCI β=0.18, p=0.07; Dementia β=0.52, p=0.02. CU, cognitively unimpaired; MCI, mild cognitive impairment; SCD, subjective cognitive decline.

### Plasma P-tau181 x APOE ε4 Carrier Status and Cognition

P-tau181 interacted with *APOE* ε4 carrier status on memory (β=-0.006, p-value=0.02) and executive functioning (β=-0.006, p-value=0.05) composite scores; however, these results did not persist after FDR correction. P-tau181 did not interact with *APOE* ε4 carrier status on language (β=-0.005, p-value=0.07), visuospatial (β=-0.007, p-value=0.58), self-reported SCD (β=0.16, p-value=0.49), or informant-reported SCD (β=0.18, p-value=0.13) scores. See **Table 4** for results stratified by *APOE* ε4 carrier status.

**Table 4.** Plasma P-Tau181 x *APOE* **ε**4 Status Interactions on Cognitive and SCD Outcomes

|  | *p-tau x APOE ε4 status* |  | *ε4 negative (n=671)* |  | *ε4 positive (n=516)* |  |
| --- | --- | --- | --- | --- | --- | --- |
|  | β | *p*-value | β | *p*-value | β | *p*-value |
| Memory | -0.006 | **0.02** | -0.004 | **0.02** | -0.01 | **<0.001\*** |
| Executive Functions | -0.006 | **0.05** | -0.002 | 0.35 | -0.008 | **<0.001\*** |
| Language | -0.005 | 0.07 | -0.002 | 0.36 | -0.006 | **0.003\*** |
| Visuospatial | -0.002 | 0.58 | -0.0004 | 0.85 | -0.002 | 0.44 |
| ECog Self | 0.16 | 0.49 | -0.06 | 0.66 | 0.18 | 0.47 |
| ECog Informant | 0.18 | 0.13 | 0.07 | 0.34 | 0.27 | **0.01\*** |

Note: Models were adjusted for age, sex, education, and cognitive diagnosis. Bold font indicates *p*-value<0.05. \*FDR-corrected *p*-value<0.05. APOE, apolipoprotein E gene; ECog, Everyday Cognition questionnaire.

### Plasma P-tau181 x Age and Cognition

P-tau181 interacted with age on memory (β=0.0004, p-value=0.005) and executive functioning (β=0.0004, p-value=0.02) composite scores, but not on other outcomes (*p*-values>0.10). Models stratified by age showed that higher p-tau181 levels were associated with lower scores on memory (β=-0.01, p-value<0.001), executive functioning (β=-0.01, p-value<0.001), and language (β=-0.007, p-value=0.001) composite scores and informant-reported SCD (β=0.17, p-value=0.05) in younger participants. Higher p-tau181 levels were also associated with worse memory performance in older participants (β=-0.006, p-value<0.001), though to a lesser degree than in younger participants. Results were comparable after FDR correction. See **Table 5** for results stratified by age group and **Figure 2** for illustrations of stratified analyses.

**Table 5.** Plasma P-Tau181 x Age Interactions on Cognitive and SCD Outcomes

|  | *p-tau x age* |  | *Age <75 years (n=594)* |  | *Age ≥75 years (n=592)* |  |
| --- | --- | --- | --- | --- | --- | --- |
|  | β | *p*-value | β | *p*-value | β | *p*-value |
| Memory | 0.0004 | **0.005\*** | -0.01 | **<0.001\*** | -0.006 | **<0.001\*** |
| Executive Functions | 0.0004 | **0.02** | -0.01 | **<0.001\*** | -0.001 | 0.47 |
| Language | 0.0001 | 0.43 | -0.007 | **0.001\*** | -0.003 | 0.12 |
| Visuospatial | 0.0002 | 0.23 | -0.004 | 0.12 | 0.001 | 0.68 |
| ECog Self | -0.03 | 0.11 | 0.22 | 0.58 | -0.02 | 0.91 |
| ECog Informant | -0.01 | 0.19 | 0.17 | **0.05** | 0.16 | 0.07 |

Note: Models were adjusted for age, sex, education, APOE ε4 status, and cognitive diagnosis. Bold font indicates *p*-value<0.05. \*FDR-corrected *p*-value<0.05. APOE, apolipoprotein E gene; ECog, Everyday Cognition questionnaire.

**Figure 2.**
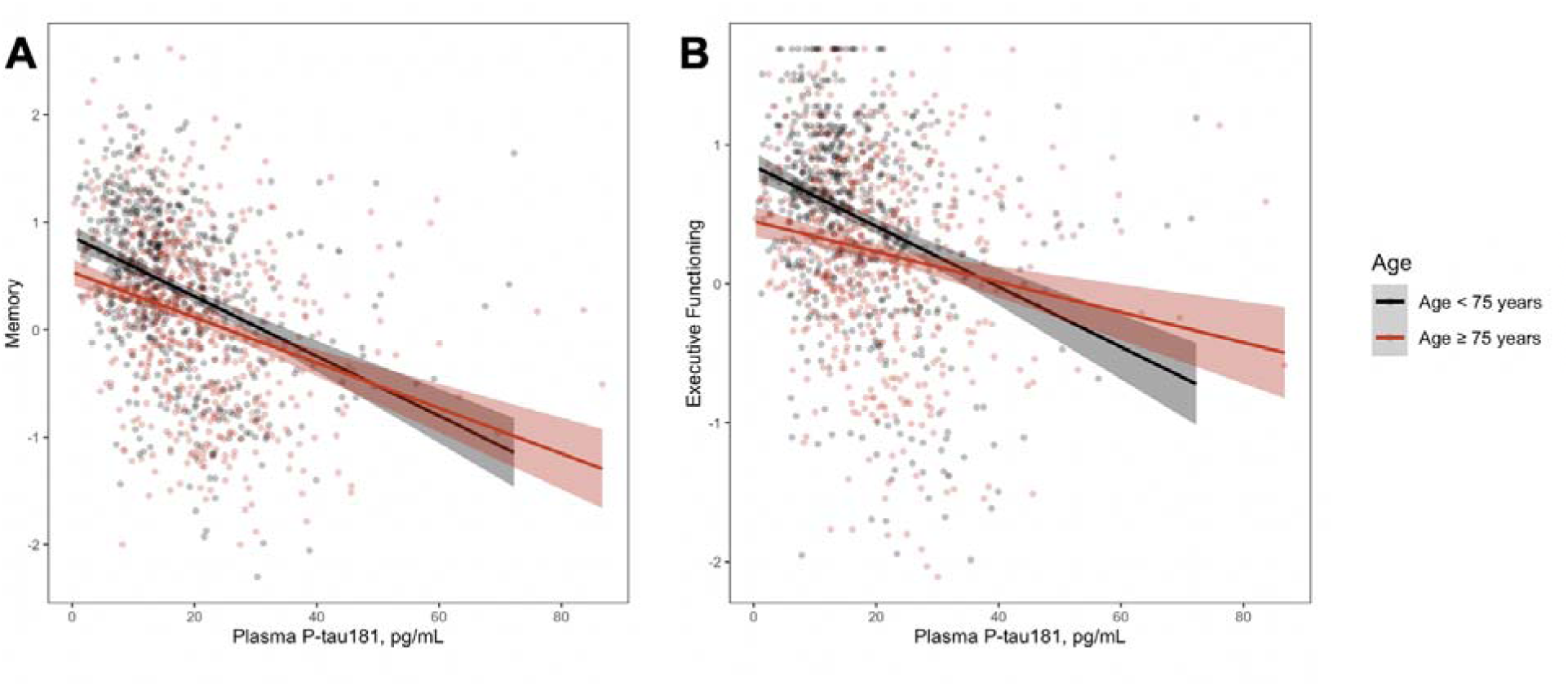
Plasma p-tau181 x Age Interactions on Cognitive Outcomes. Lines reflect cognitive domain scores corresponding to plasma p-tau181 levels. Shading reflects 95% confidence interval. (A) Associations between plasma p-tau181 levels and memory composite score, stratified by age group; <75 years β=-0.01, p<0.001; ≥75 years β=-0.006, p<0.001. (B) Associations between plasma p-tau181 levels and executive functioning composite score, stratified by age group; <75 years β=-0.01, p<0.001; ≥75 years β=-0.001, p=0.47.

### Plasma P-tau181 x Sex and Cognition

P-tau181 interacted with sex on memory composite score (β=-0.006, p-value=0.03) and self-reported SCD scores (β=-0.68, p-value=0.02). However, these results were attenuated after FDR correction (*p*-values>0.06). In models stratified by sex, higher p-tau181 levels were associated with lower memory in both men (β=-0.006, p-value=0.002) and women (β=-0.009, p-value=0.0001), with stronger associations in women. Higher p-tau181 levels were also associated with greater informant-reported SCD scores in men (β=0.23, p-value=0.005), but not women (p-value=0.46), and with executive function composite scores in women (β=-0.006, p-value=0.01), but not in men (p-value=0.06). All stratified results persisted after FDR correction. See **Table 6** for results stratified by sex and **Figure 3** for illustrations of stratified analyses.

**Table 6.** Plasma P-Tau181 x Sex Interactions on Cognitive and SCD Outcomes

|  | *p-tau x sex* |  | Male (n=644) |  | Female (n=543) |  |
| --- | --- | --- | --- | --- | --- | --- |
| | $\beta$ | *p*-value | $\beta$ | *p*-value | $\beta$ | *p*-value |
| Memory | -0.006 | **0.03** | -0.006 | **0.002\*** | -0.009 | **0.0001\*** |
| Executive Functioning | -0.002 | 0.48 | -0.004 | 0.06 | -0.006 | **0.01\*** |
| Language | -0.0001 | 0.97 | -0.004 | 0.06 | -0.004 | 0.07 |
| Visuospatial | 0.0002 | 0.95 | -0.0009 | 0.63 | -0.002 | 0.50 |
| ECog Self | -0.68 | **0.02** | 0.20 | 0.18 | -0.46 | 0.07 |
| ECog Informant | -0.07 | 0.57 | 0.23 | **0.005\*** | 0.07 | 0.46 |

Note: Models were adjusted for age, education, *APOE* ε4 status, and cognitive diagnosis. Bold font indicates *p*-value<0.05. \*FDR-corrected *p*-value<0.05. *APOE*, apolipoprotein E gene; ECog, Everyday Cognition questionnaire.

**Figure 3.**
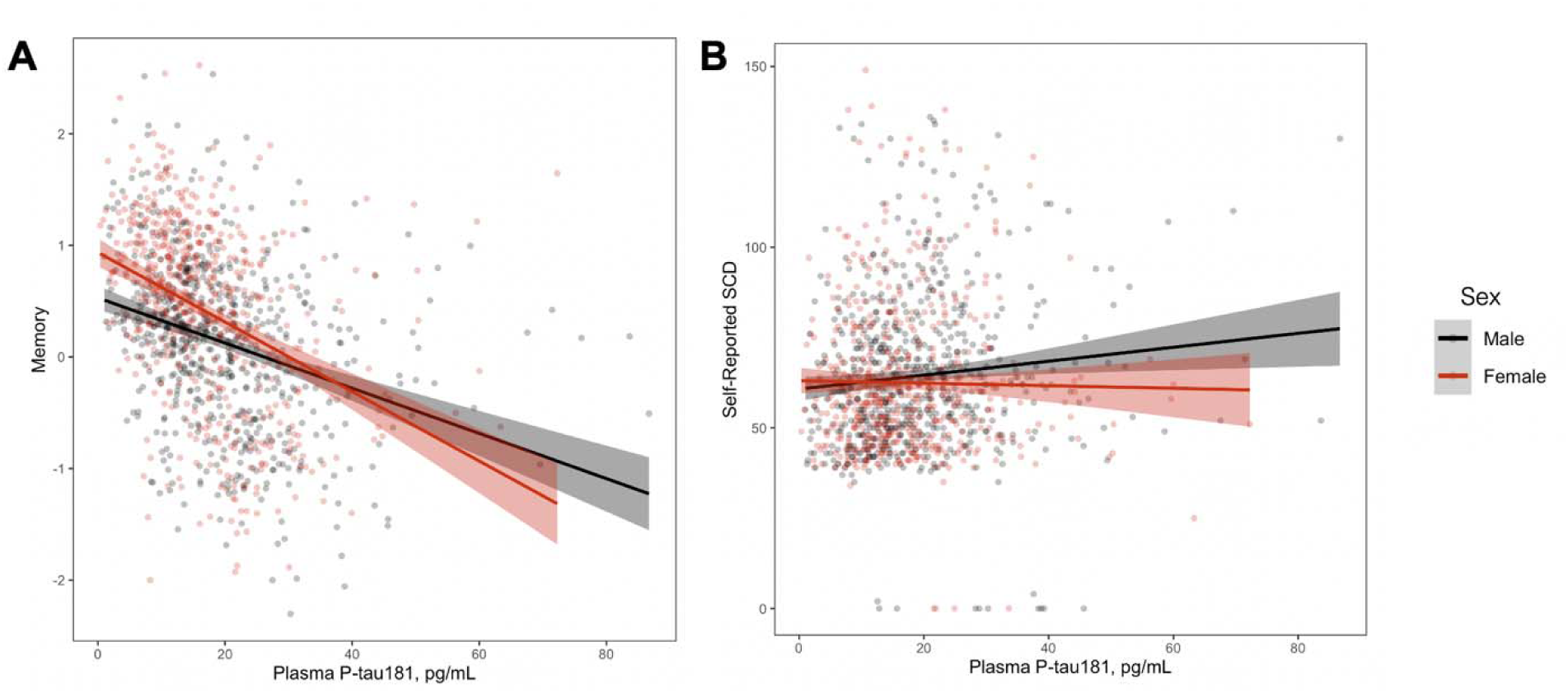
Plasma p-tau181 x Sex Interactions on Cognitive Outcomes. Lines reflect cognitive domain and self-reported SCD scores corresponding to plasma p-tau181 levels. Shading reflects 95% confidence interval. (A) Associations between plasma p-tau181 levels and memory composite score, stratified by sex; male β=-0.006, p=0.002; female β=-0.009, p=0.0001. (B) Associations between plasma p-tau181 levels and self-reported SCD score, stratified by sex; male β=0.20, p=0.18; female β=-0.46, p=0.07. SCD, subjective cognitive decline.

### Sensitivity Analyses

Results across all analyses were largely unchanged after excluding outliers and using mean scores, rather than total scores, for all SCD analyses.

## Discussion

Among community-dwelling older adults across the AD clinical spectrum, higher levels of plasma p-tau181 were associated with lower cognitive performance in the domains of memory, executive functioning, and language, and greater informant-reported SCD. Visuospatial abilities and self-report SCD were not associated with plasma p-tau181. Results were most prominent in individuals who were diagnosed with MCI or dementia, *APOE* ε4 carriers, women, and younger in age. Plasma p-tau181 was not associated with objective or subjective cognition in CU individuals.

The observed associations between plasma p-tau181 and cognition align with past work in support of plasma p-tau181 as a marker of AD neurodegeneration.^4^ Plasma p-tau181 was robustly associated with episodic memory functioning, the earliest and most typical cognitive deficit seen in AD resulting from the formation of neurofibrillary tangles and subsequent atrophy in memory-critical structures of the medial temporal lobes. Additionally, aligning with recent published work,^35^ plasma p-tau181 was associated with language and executive functioning, cognitive domains that become impaired due to evolving neurodegeneration in the frontal and temporal association cortices secondary to pathological progression of AD.^36^ These patterns were strongest in individuals with MCI or dementia and *APOE* ε4 carriers. The diagnostic association is likely due to the fact that people in preclinical or dementia states of AD evidence the highest levels of plasma p-tau181^1^ and lowest levels of cognition, and the current results lend further support to the validity of plasma p-tau181 as a clinical marker of AD. Of note, p-tau181 was not associated with memory in dementia likely due to the ubiquity of memory problems at this stage of cognitive impairment and plateauing levels of p-tau181 in advanced disease.^2^ Similarly, *APOE* ε4 is associated with worse memory and reasoning in older adults,^37^ as well as a higher accumulation rate of tau in AD.^38^ As such, *APOE* ε4 carriers can be more prone to neurodegeneration and current results suggest that *APOE* genotype may be an important consideration when understanding how this plasma biomarker predicts who may display or develop cognitive impairment.

The lack of association between plasma p-tau181 and cognition in CU individuals was somewhat surprising given that plasma p-tau181 elevates early in the disease process, purportedly shortly after abnormal amyloid-β accumulation.^2^ Multiple factors may be contributing to the observed findings. One, it is possible that while plasma p-tau181 can be elevated early in the disease course, the variance of either the biomarker or cognitive performances observed in this CU sample was not sufficient to observe differences. As discussed elsewhere,^39^ the selection criteria for the ADNI cohorts limits the presence of comorbidities or alternative etiologies (e.g., cerebrovascular disease), thus the sample here is likely healthier than the general population. With fewer medical comorbidities, CU individuals may be less likely to have other non-AD pathologies such as cerebrovascular disease, and thus may be more resilient to early pathological changes. Second, the cognitive outcomes may lack sensitivity in identifying very early and subtle cognitive changes. Education confounds neuropsychological test performance and may protect against cognitive changes related to the earliest accumulation of AD pathology.^40^ In a highly educated cohort, such as this one, AD pathological changes may not produce a measurable cognitive deficit until later in the disease progression.

As expected, informant-reported SCD was associated with plasma p-tau181 within the MCI and dementia phases of AD, while self-reported SCD was not associated. This pattern is likely secondary to the presence of anosognosia, a common clinical feature of Alzheimer’s disease, that can occur prior to frank dementia.^41^ Self-reported SCD was unrelated to plasma p-tau181, even in cognitively unimpaired participants. This finding was somewhat unexpected as self-reported SCD has been linked with amyloid deposition, and previous findings suggest that p-tau181 elevations shortly follow increased amyloid burden.^42^ This lack of association could indicate that self-reported SCD occurs prior to detectable changes in plasma p-tau181. Alternatively, self-reported SCD is present due to a multitude of factors and the null findings may be related to other etiologies driving the subjective cognitive weaknesses (e.g., mood and personality factors, learning disability).

Age and sex both appeared to modify associations between plasma p-tau181 and cognition, with stronger findings in younger participants (age <75 years) and women, likely due to multiple factors. First, with increasing age comes increasing risk of developing other neuropathologies that contribute to cognitive changes.^43^ Additionally, younger age of AD onset is associated with a more aggressive disease course, greater pathological burden, and faster clinical decline.^44^ Similar considerations likely occur with sex. Women may be more clinically susceptible to pathological changes of AD. For example, when comparing women and men with similar levels of AD pathology, women show greater problems with cognition and across more domains as compared to men.^45^ Thus, younger participants or women may be more likely to have greater cognitive impairment across more domains in the presence of AD pathology.

One unexpected finding was that, despite plasma p-tau181 being more strongly associated with cognition in women, it was associated with greater informant-reported SCD in men, but not women. These findings are previously unreported to our knowledge and could suggest that informant-reported SCD in men may be more reflective of pathological changes due to AD as compared to women. These results could be related to women being more likely to experience cognitive changes that are due to alternative etiologies (e.g., depression) that are not significant enough to produce objective changes on neuropsychological measures but are producing changes that are noticeable to an informant. Taken together, these results suggest that plasma p-tau181 is more strongly linked with objective cognition in women but subjective cognition in men.

This study has notable strengths including utilizing a comprehensive neuropsychological assessment with more granular measures of objective cognition that have been developed using sophisticated statistical techniques. The current study included measures of self-and informant report SCD, an early marker of increased risk for cognitive decline. Further, the large sample size in this study allows for examination of numerous potentially modifying factors, including cognitive diagnosis, *APOE* ε4 carrier status, age, and sex. Limitations include a lack of diversity in important factors, such as race/ethnicity and education in ADNI, thereby limiting generalizability. Further, neuropsychological composite scores, while advantageous in that they reduce the risk of Type I error related to multiple comparisons, may have less sensitivity in preclinical disease states as compared to certain individual neuropsychological measures or process scores.

In sum, we found that plasma p-tau181 correlates with clinical changes that are typical of AD, thereby providing new evidence consistent with past work showing that this novel biomarker is useful in detecting clinical changes specific to AD pathology. Further, we highlighted the specific demographic, diagnostic, and clinical factors which modify the association between plasma p-tau181 and cognition. Plasma p-tau181, assessed cross-sectionally, may be most clinically useful in individuals with some degree of objective cognitive impairment, though *APOE* ε4 carrier status, age, and sex should be considered when understanding these associations.

## Funding and Acknowledgements

Funding to support this work includes: K23-AG045966 (KAG), R01-AG062826 (KAG), R01-AG073439 (LCD), F32-AG076276 (CJB), K24-AG046373 (ALJ), T32-AG058524 (CJB). HZ is a Wallenberg Scholar supported by grants from the Swedish Research Council (#2022-01018 and #2019-02397), the European Union’s Horizon Europe research and innovation programme under grant agreement No 101053962, Swedish State Support for Clinical Research (#ALFGBG-71320), the Alzheimer Drug Discovery Foundation (ADDF), USA (#201809-2016862), the AD Strategic Fund and the Alzheimer’s Association (#ADSF-21-831376-C, #ADSF-21-831381-C, and #ADSF-21-831377-C), the Bluefield Project, the Olav Thon Foundation, the Erling-Persson Family Foundation, Stiftelsen för Gamla Tjänarinnor, Hjärnfonden, Sweden (#FO2022-0270), the European Union’s Horizon 2020 research and innovation programme under the Marie Skłodowska-Curie grant agreement No 860197 (MIRIADE), the European Union Joint Programme – Neurodegenerative Disease Research (JPND2021-00694), the National Institute for Health and Care Research University College London Hospitals Biomedical Research Centre, and the UK Dementia Research Institute at UCL (UKDRI-1003).

Data collection and sharing for this project was funded by the Alzheimer’s Disease Neuroimaging Initiative (ADNI) (National Institutes of Health Grant U01 AG024904) and DOD ADNI (Department of Defense award number W81XWH-12-2-0012). ADNI is funded by the National Institute on Aging, the National Institute of Biomedical Imaging and Bioengineering, and through generous contributions from the following: AbbVie, Alzheimer’s Association; Alzheimer’s Drug Discovery Foundation; Araclon Biotech; BioClinica, Inc.; Biogen; Bristol-Myers Squibb Company; CereSpir, Inc.; Cogstate; Eisai Inc.; Elan Pharmaceuticals, Inc.; Eli Lilly and Company; EuroImmun; F. Hoffmann-La Roche Ltd and its affiliated company Genentech, Inc.; Fujirebio; GE Healthcare; IXICO Ltd.; Janssen Alzheimer Immunotherapy Research & Development, LLC.; Johnson & Johnson Pharmaceutical Research & Development LLC.; Lumosity; Lundbeck; Merck & Co., Inc.; Meso Scale Diagnostics, LLC.; NeuroRx Research; Neurotrack Technologies; Novartis Pharmaceuticals Corporation; Pfizer Inc.; Piramal Imaging; Servier; Takeda Pharmaceutical Company; and Transition Therapeutics. The Canadian Institutes of Health Research is providing funds to support ADNI clinical sites in Canada. Private sector contributions are facilitated by the Foundation for the National Institutes of Health (www.fnih.org). The grantee organization is the Northern California Institute for Research and Education, and the study is coordinated by the Alzheimer’s Therapeutic Research Institute at the University of Southern California. ADNI data are disseminated by the Laboratory for Neuro Imaging at the University of Southern California.

## Data Availability

All data produced in the present study are available upon reasonable request to the authors

https://www.vmacdata.org

## References

1. Janelidze S, Mattsson N, Palmqvist S, et al. Plasma P-tau181 in Alzheimer’s disease: relationship to other biomarkers, differential diagnosis, neuropathology and longitudinal progression to Alzheimer’s dementia. Nat Med. 2020;26(3):379–386. doi:10.1038/s41591-020-0755-1

2. Moscoso A, Grothe MJ, Ashton NJ, et al. Time course of phosphorylated-tau181 in blood across the Alzheimer’s disease spectrum. Brain. 2021;144(1):325–339. doi:10.1093/brain/awaa399

3. Karikari TK, Pascoal TA, Ashton NJ, et al. Blood phosphorylated tau 181 as a biomarker for Alzheimer’s disease: a diagnostic performance and prediction modelling study using data from four prospective cohorts. Lancet Neurol. 2020;19(5):422–433. doi:10.1016/S1474-4422(20)30071-5

4. Moscoso A, Grothe MJ, Ashton NJ, et al. Longitudinal Associations of Blood Phosphorylated Tau181 and Neurofilament Light Chain With Neurodegeneration in Alzheimer Disease. JAMA Neurol. Published online January 11, 2021. doi:10.1001/jamaneurol.2020.4986

5. Blennow K. Plasma P-tau immunoassays, methodological aspects and clinical performance. Alzheimers Dement. 2020;16(S5):e037515. doi:10.1002/alz.037515

6. Sutterer MJ, Tranel D. Neuropsychology and cognitive neuroscience in the fMRI era: A recapitulation of localizationist and connectionist views. Neuropsychology. 2017;31(8):972–980. doi:10.1037/neu0000408

7. Wisdom NM, Mignogna J, Collins RL. Variability in Wechsler Adult Intelligence Scale-IV subtest performance across age. Arch Clin Neuropsychol Off J Natl Acad Neuropsychol. 2012;27(4):389–397. doi:10.1093/arclin/acs041

8. McCarrey AC, An Y, Kitner-Triolo MH, Ferrucci L, Resnick SM. Sex differences in cognitive trajectories in clinically normal older adults. Psychol Aging. 2016;31(2):166–175. doi:10.1037/pag0000070

9. Rawle MJ, Davis D, Bendayan R, Wong A, Kuh D, Richards M. Apolipoprotein-E (Apoe) ε4 and cognitive decline over the adult life course. Transl Psychiatry. 2018;8(1):18. doi:10.1038/s41398-017-0064-8

10. Weintraub S, Wicklund AH, Salmon DP. The Neuropsychological Profile of Alzheimer Disease. Cold Spring Harb Perspect Med. 2012;2(4):a006171. doi:10.1101/cshperspect.a006171

11. Hachinski VC, Iliff LD, Zilhka E, et al. Cerebral Blood Flow in Dementia. Arch Neurol. 1975;32(9):632–637. doi:10.1001/archneur.1975.00490510088009

12. Yesavage JA, Brink TL, Rose TL, et al. Development and validation of a geriatric depression screening scale: a preliminary report. J Psychiatr Res. 1982;17(1):37–49. doi:10.1016/0022-3956(82)90033-4

13. Wechsler D. Wechsler Memory Scale-Revised Manual. The Psychological Corporation; 1987.

14. Folstein MF, Folstein SE, McHugh PR. “Mini-mental state”: A practical method for grading the cognitive state of patients for the clinician. J Psychiatr Res. 1975;12(3):189–198. doi:10.1016/0022-3956(75)90026-6

15. Hughes CP, Berg L, Danziger W, Coben LA, Martin RL. A New Clinical Scale for the Staging of Dementia. Br J Psychiatry. 1982;140(6):566–572. doi:10.1192/bjp.140.6.566

16. McKhann G, Drachman D, Folstein M, Katzman R, Price D, Stadlan EM. Clinical diagnosis of Alzheimer’s disease: report of the NINCDS-ADRDA Work Group under the auspices of Department of Health and Human Services Task Force on Alzheimer’s Disease. Neurology. 1984;34(7):939–944.

17. Crane PK, Carle A, Gibbons LE, et al. Development and assessment of a composite score for memory in the Alzheimer’s Disease Neuroimaging Initiative (ADNI). Brain Imaging Behav. 2012;6(4):502–516. doi:10.1007/s11682-012-9186-z

18. Rey A. L’examen Clinique En Psychologie. [The Clinical Examination in Psychology.]. Presses Universitaries De France; 1958:222.

19. Rosen WG, Mohs RC, Davis KL. A new rating scale for Alzheimer’s disease. Am J Psychiatry. 1984;141(11):1356–1364. doi:10.1176/ajp.141.11.1356

20. Gibbons LE, Carle AC, Mackin RS, et al. A composite score for executive functioning, validated in Alzheimer’s Disease Neuroimaging Initiative (ADNI) participants with baseline mild cognitive impairment. Brain Imaging Behav. 2012;6(4):517–527. doi:10.1007/s11682-012-9176-1

21. Weintraub S, Salmon D, Mercaldo N, et al. The Alzheimer’s Disease Centers’ Uniform Data Set (UDS): The Neuropsychological Test Battery. Alzheimer Dis Assoc Disord. 2009;23(2):91–101. doi:10.1097/WAD.0b013e318191c7dd

22. Reitan RM. Trail Making Test: Manual for Administration and Scoring. Reitan Neuropsychology Laboratory; 1992.

23. Wechsler A. Wechsler Adult Intelligence Scale-Revised. Psychological Corporation; 1987.

24. Sunderland T, Hill JL, Mellow AM, et al. Clock drawing in Alzheimer’s disease. A novel measure of dementia severity. J Am Geriatr Soc. 1989;37(8):725–729.

25. Choi S, Mukherjee S, Gibbons LE, et al. Development and validation of language and visuospatial composite scores in ADNI. Alzheimers Dement Transl Res Clin Interv. 2020;6(1):e12072. doi:10.1002/trc2.12072

26. Kaplan E, Goodglass H, Weintraub S. Boston Naming Test. PsycTESTS. Published online 1983. doi:10.1037/t27208-000

27. Nasreddine ZS, Phillips NA, Bédirian V, et al. The Montreal Cognitive Assessment, MoCA: A Brief Screening Tool For Mild Cognitive Impairment. J Am Geriatr Soc. 2005;53(4):695–699. doi:10.1111/j.1532-5415.2005.53221.x

28. Farias ST, Mungas D, Reed BR, et al. The measurement of everyday cognition (ECog): scale development and psychometric properties. Neuropsychology. 2008;22(4):531–544. doi:10.1037/0894-4105.22.4.531

29. Salthouse TA. When does age-related cognitive decline begin? Neurobiol Aging. 2009;30(4):507–514. doi:10.1016/j.neurobiolaging.2008.09.023

30. Levine DA, Gross AL, Briceño EM, et al. Sex Differences in Cognitive Decline Among US Adults. JAMA Netw Open. 2021;4(2):e210169. doi:10.1001/jamanetworkopen.2021.0169

31. Alley D, Suthers K, Crimmins E. Education and Cognitive Decline in Older Americans: Results From the AHEAD Sample. Res Aging. 2007;29(1):73–94. doi:10.1177/0164027506294245

32. Caselli RJ, Chen K, Locke DEC, et al. Subjective cognitive decline: Self and informant comparisons. Alzheimers Dement J Alzheimers Assoc. 2014;10(1):93–98. doi:10.1016/j.jalz.2013.01.003

33. Bretsky P, Guralnik JM, Launer L, Albert M, Seeman TE, MacArthur Studies of Successful Aging. The role of APOE-epsilon4 in longitudinal cognitive decline: MacArthur Studies of Successful Aging. Neurology. 2003;60(7):1077–1081. doi:10.1212/01.wnl.0000055875.26908.24

34. Benjamini Y, Hochberg Y. Controlling the False Discovery Rate: A Practical and Powerful Approach to Multiple Testing. J R Stat Soc Ser B Methodol. 1995;57(1):289–300.

35. Wang YL, Chen J, Du ZL, et al. Plasma p-tau181 Level Predicts Neurodegeneration and Progression to Alzheimer’s Dementia: A Longitudinal Study. Front Neurol. 2021;12:695696. doi:10.3389/fneur.2021.695696

36. Braak H, Braak E. Neuropathological stageing of Alzheimer-related changes. Acta Neuropathol (Berl). 1991;82(4):239–259.

37. Gharbi-Meliani A, Dugravot A, Sabia S, et al. The association of APOE ε4 with cognitive function over the adult life course and incidence of dementia: 20Lyears follow-up of the Whitehall II study. Alzheimers Res Ther. 2021;13(1):5. doi:10.1186/s13195-020-00740-0

38. Baek MS, Cho H, Lee HS, Lee JH, Ryu YH, Lyoo CH. Effect of APOE ε4 genotype on amyloid-β and tau accumulation in Alzheimer’s disease. Alzheimers Res Ther. 2020;12:140. doi:10.1186/s13195-020-00710-6

39. Thomas KR, Weigand AJ, Cota IH, et al. Intrusion errors moderate the relationship between blood glucose and regional cerebral blood flow in cognitively unimpaired older adults. Brain Imaging Behav. 2022;16(1):219–227. doi:10.1007/s11682-021-00495-8

40. Knopman DS, Caselli RJ. Appraisal of cognition in preclinical Alzheimer’s disease: a conceptual review. Neurodegener Dis Manag. 2012;2(2):183–195. doi:10.2217/NMT.12.5

41. Gerretsen P, Chung JK, Shah P, et al. Anosognosia Is an Independent Predictor of Conversion From Mild Cognitive Impairment to Alzheimer’s Disease and Is Associated With Reduced Brain Metabolism. J Clin Psychiatry. 2017;78(9):805. doi:10.4088/JCP.16m11367

42. Barthélemy NR, Li Y, Joseph-Mathurin N, et al. A soluble phosphorylated tau signature links tau, amyloid and the evolution of stages of dominantly inherited Alzheimer’s disease. Nat Med. 2020;26(3):398–407. doi:10.1038/s41591-020-0781-z

43. Robinson JL, Lee EB, Xie SX, et al. Neurodegenerative disease concomitant proteinopathies are prevalent, age-related and APOE4-associated. Brain. 2018;141(7):2181–2193. doi:10.1093/brain/awy146

44. Wattmo C, Wallin ÅK. Early-versus Late-Onset Alzheimer Disease: Long-Term Functional Outcomes, Nursing Home Placement, and Risk Factors for Rate of Progression. Dement Geriatr Cogn Disord EXTRA. 2017;7(1):172–187. doi:10.1159/000455943

45. Laws KR, Irvine K, Gale TM. Sex differences in cognitive impairment in Alzheimer’s disease. World J Psychiatry. 2016;6(1):54-65. doi:10.5498/wjp.v6.i1.54

